# Trends and clinical characteristics of COVID-19 vaccine recipients: a federated analysis of 57.9 million patients’ primary care records in situ using OpenSAFELY

**DOI:** 10.1101/2021.01.25.21250356

**Authors:** The OpenSAFELY Collaborative, Helen J Curtis, Peter Inglesby, Caroline E Morton, Brian MacKenna, Alex J Walker, Jessica Morley, Amir Mehrkar, Seb Bacon, George Hickman, Chris Bates, Richard Croker, David Evans, Tom Ward, Jonathan Cockburn, Simon Davy, Krishnan Bhaskaran, Anna Schultze, Christopher T Rentsch, Elizabeth Williamson, William Hulme, Amelia Green, Anna Rowan, Louis Fisher, Helen I McDonald, Laurie Tomlinson, Rohini Mathur, Henry Drysdale, Rosalind M Eggo, Kevin Wing, Angel YS Wong, Harriet Forbes, John Parry, Frank Hester, Sam Harper, Shaun O’Hanlon, Alex Eavis, Richard Jarvis, Dima Avramov, Paul Griffiths, Aaron Fowles, Nasreen Parkes, Ian J Douglas, Stephen JW Evans, Liam Smeeth, Ben Goldacre

## Abstract

**Background:** On December 8th 2020, NHS England administered the first COVID-19 vaccination as part of an ambitious vaccination programme during a global health emergency.

**Aims:** To describe trends and variation in vaccine coverage by key clinical and demographic groups; to create a framework for near-real-time monitoring of vaccine coverage in key subgroups.

**Methods:** Working on behalf of NHS England we analysed 57.9 million patient records in situ and in near-real-time within the infrastructure of the Electronic Health Record (EHR) software vendors EMIS and TPP using OpenSAFELY. We describe vaccine coverage and time trends across a range of demographic and fine-grained clinical subgroups in eight Joint Committee on Vaccination and Immunisation (JCVI) priority cohorts.

**Results:** 20,852,692 patients (36%) received a COVID-19 vaccine between December 8th 2020 and March 17th 2021. Of patients aged ≥80 not in a care home (JCVI group 2) 94.7% received a vaccine, but with substantial variation by ethnicity (White 96.2% vaccinated, Black 68.3%) and deprivation (least deprived 96.6%, most deprived 90.7%). Overall, patients with pre-existing medical conditions were equally or more likely to be vaccinated with two exceptions: severe mental illness (89.5% vaccinated) and learning disability (91.4%). 275,205 vaccine recipients were identified as care home residents (priority group 1; 91.2% coverage). 1,257,914 (6.0%) recipients have had a second dose. Detailed characteristics of recipients in all cohorts are reported.

**Conclusions:** The NHS in England has rapidly delivered mass vaccination. We were able to deploy a data monitoring framework using publicly auditable methods and a secure, in-situ processing model, using linked but pseudonymised patient-level NHS data on 57.9 million patients with very short delays from vaccine administration to completed analysis. Targeted activity may be needed to address lower vaccination coverage observed among certain key groups: ethnic minorities, those living in deprived areas, and people with severe mental illness or learning disabilities.

## Background

On December 8th 2020, the NHS in England administered the first COVID-19 vaccination as part of an ambitious vaccine programme to combat the ongoing pandemic due to severe acute respiratory syndrome coronavirus 2 (SARS-CoV-2). Vaccination is one of the most cost-effective ways of avoiding disease, and worldwide vaccinations prevent 2-3 million deaths per year, but new vaccines can take many years or decades to develop and become part of routine practice [1]. Since the outset of the COVID-19 pandemic, teams of scientists, clinical trialists and regulators around the world have worked at unprecedented speed, with more than 200 vaccines currently being tested [2]. The UK medicines regulator approved two COVID-19 vaccines for use before the end of 2020: the *Pfizer-BioNTech* mRNA vaccine and the *AstraZeneca-Oxford* vaccine. The *Moderna* vaccine was subsequently approved on January 8th and more vaccine approvals are expected in 2021 [3,4].

For the UK the independent Joint Committee on Vaccination and Immunisation (JCVI) provides recommendations on vaccinations to the government and England’s National Health Service (NHS). In early December 2020 the JCVI recommended nine priority groups for vaccination (box 1), largely based on risk of death from COVID-19.[5] This was the basis for the NHS England vaccination programme, with due recognition that vaccination of care home residents may initially lag behind other groups due to complexities around storing and distributing *Pfizer-BioNTech* outside of larger healthcare settings without appropriate cold chain facilities [6]. Vaccinations were administered initially in hospitals and a number of primary care centres; they are now being administered in GP surgeries, community pharmacies and newly established mass vaccination centres. Both vaccines administered to date require two doses. Given high infection rates and the relatively high protection thought to be offered by the first dose, after the start of the campaign the JCVI recommended extending the interval to 12 weeks. This strategy was intended to prevent the most deaths and hospitalisations through maximising the number of patients with some protection against the virus as quickly as possible, although GPs were initially allowed some discretion in the exact timing of the second dose [7,8].

OpenSAFELY is a new secure analytics platform for electronic patient records built by our group on behalf of NHS England to deliver urgent academic and operational research during the pandemic [9,10]. Analyses run across all patients’ full coded pseudonymised primary care records, and includes 95% of people registered with an English general practice, those where EMIS or TPP electronic health record (EHR) software is deployed, with patient-level linkage to various sources, such as secondary care data. Code and analysis is shared openly for inspection and re-use. Vaccine administration details are recorded in the National Immunisation Management Service (NIMS) and electronically transmitted to every individual’s GP record on a daily basis.

We therefore set out to: assess the coverage of COVID-19 vaccination in all patients registered with TPP and EMIS practices in England in near real-time; and to describe how coverage varied between key clinical and demographic subgroups.

Box 1 – *Priority groups for vaccination advised by the Joint Committee on Vaccination and Immunisation [5]*

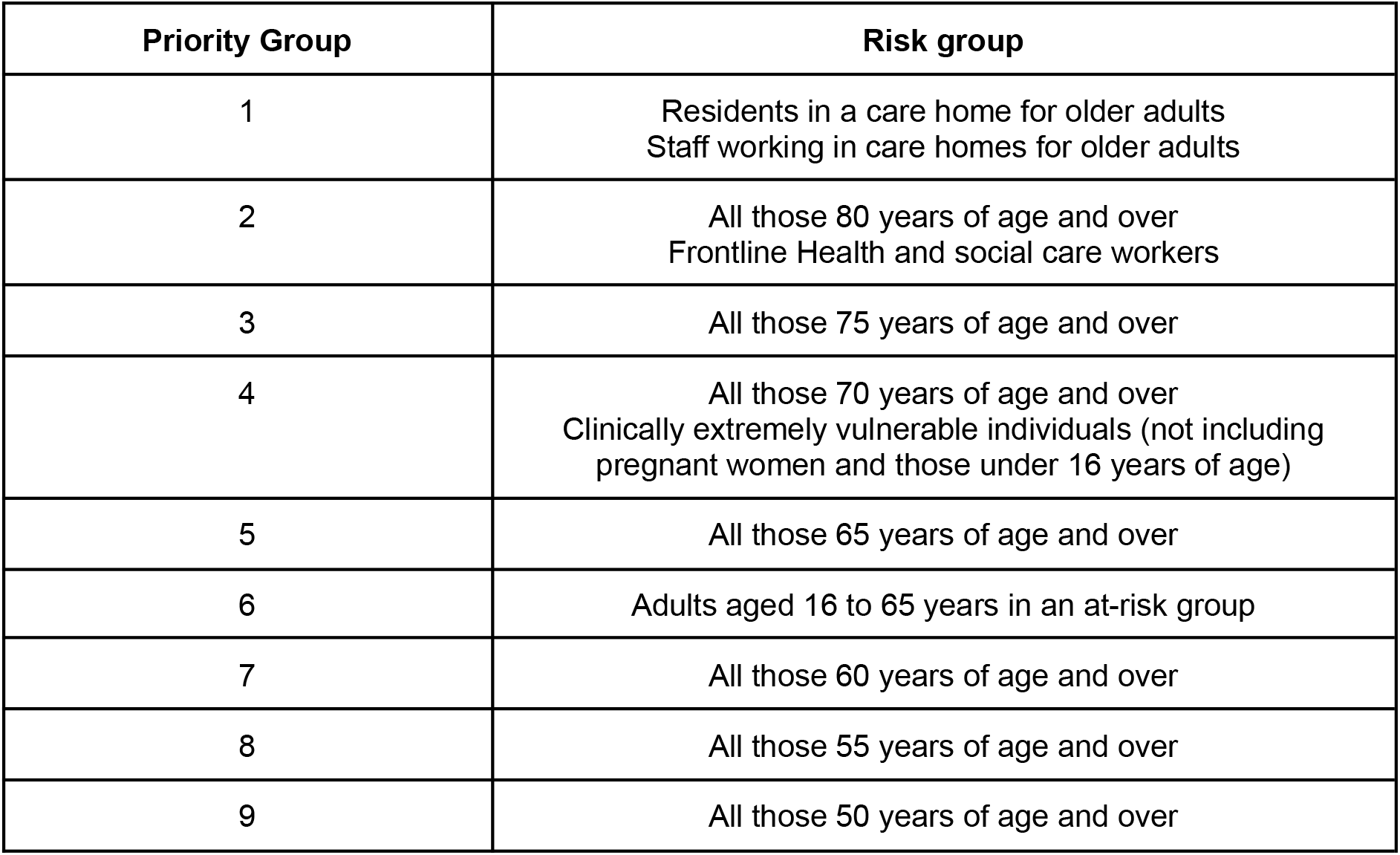

## Methods

### Study design

We conducted a retrospective cohort study using general practice primary care electronic health record data from all England GP practices supplied by the electronic health record vendors EMIS and TPP. The cohort study began on 8th December 2020, the start of the national vaccination campaign, and ended on March 25th. We are producing weekly vaccine coverage reports with a subset of this data [11] and will update this analysis regularly with extended follow-up time using near real time data as the vaccination campaign progresses.

### Data Source

Primary care records managed by the GP software providers EMIS and TPP were accessed through OpenSAFELY, an open source data analytics platform created by our team on behalf of NHS England to address urgent COVID-19 research questions (https://opensafely.org). OpenSAFELY provides a secure software interface allowing a federated analysis of pseudonymized primary care patient records from England in near real-time within the EMIS and TPP highly secure data environments. Non-disclosive, aggregated results are exported to GitHub where further data processing and analysis takes place. This avoids the need for large volumes of potentially disclosive pseudonymised patient data to be transferred off-site. This, in addition to other technical and organisational controls, minimizes any risk of re-identification. The dataset available to the platform includes pseudonymised data such as coded diagnoses, medications and physiological parameters. No free text data are included. All activity on the platform is publicly logged and all analytic code and supporting clinical coding lists are automatically published. In addition, the framework provides assurance that the analysis is reproducible and reusable. Further details on our information governance and platform can be found below under information governance and ethics.

### Study population

We included all patients registered with a general practice using EMIS (n= 33,873,987) or TPP (n=24,056,480) in England on March 25th. We excluded patients with unknown date of birth (i.e. default age >=121) or unknown sex.

### Priority groups for vaccination

We classified patients into their JCVI priority group (box 1) using SNOMED-CT codelists and logic defined in the national *COVID-19 Vaccination Uptake Reporting Specification* developed by PRIMIS [12]. Patients were assigned only to their highest priority group and not included again as part of any other priority group. For example a 76-year-old living in a care home would be assigned to group 1 (care home residents) but not to group 3 (75-79). We did not assess eligibility as defined by occupation, i.e. health and care staff for the relevant priority groups (1 and 2) because this information is largely missing from GP records and where present, is unreliable. These patients were therefore either classified into a lower priority group where applicable (e.g. by clinical conditions or age), or as “other”, if a person did not fall into one of the nine defined groups. In line with the national reporting specification, most criteria were ascertained using the latest available data at the time of analysis, with the exception of age which was calculated as at 31 March 2021 as recommended by Public Health England.

### COVID-19 vaccine status

Vaccination information is transmitted back to patients’ primary care records in the days following vaccine administration in a designated centre. We ascertained which patients had any recorded COVID-19 vaccine administration code in their primary care record (only *Pfizer-BioNTech* mRNA vaccine or *AstraZeneca-Oxford* vaccine were available at the time of analysis). We included those vaccinated up to March 17th 2021, the latest available date of vaccinations recorded in the most recent comparable OpenSAFELY-EMIS and OpenSAFELY-TPP database build. We classed any COVID-19 vaccination within 19 days of the first dose as a duplicate record entry, and the first vaccination after this date as the second dose of the schedule.

### Key demographic and clinical characteristics of vaccinated groups

We extracted all patient demographics defined by the national reporting specification (for example, ethnicity). We also extracted demographics not defined by the specification, including the level of deprivation. Deprivation was measured by the Index of Multiple Deprivation (IMD, in quintiles, with higher values indicating greater deprivation), derived from the patient’s postcode at Lower Super Output Area.

We also describe the population according to the presence or absence of various pre-existing health problems: chronic cardiac disease; diabetes; chronic kidney disease; severe mental illness; learning disabilities; chronic neurological disease (including stroke); asplenia; morbid obesity; chronic liver disease; chronic respiratory disease; and immunosuppression.

### Codelists and implementation

Information on all characteristics were obtained from primary care records by searching TPP SystmOne and EMIS records for specific coded data. EMIS and TPP SystmOne are fully compliant with the mandated NHS standard of SNOMED-CT clinical terminology. Medicines are entered or prescribed in a format compliant with the NHS Dictionary of Medicines and Devices (dm+d) [13]. Codelists and logic for most features in the national reporting specification were automatically converted to software (https://codelists.opensafely.org/codelist/primis-covid19-vacc-uptake/).

### Analysis

We generated charts of vaccine coverage for all underlying conditions and medicines. Those not presented in this manuscript are available online for inspection in the associated GitHub repository [14].

### Software and Reproducibility

Data management and analysis was performed using the OpenSAFELY software libraries and Jupyter notebooks, both implemented using Python 3. More details are available in the Supplementary Materials. This is the first analysis delivered using federated analysis through the OpenSAFELY platform: codelists and code for data management and data analysis were specified once using the OpenSAFELY tools; then transmitted securely from the OpenSAFELY jobs server to the OpenSAFELY-TPP platform within TPP’s secure environment, and separately to the OpenSAFELY-EMIS platform within EMIS’s secure environment, where they were each executed separately against local patient data; summary results were then reviewed for disclosiveness, released, and combined for the final outputs. All code for the OpenSAFELY platform for data management, analysis and secure code execution is shared for review and re-use under open licenses at GitHub.com/OpenSAFELY. All code for data management and analysis for this paper is shared for scientific review and re-use under open licenses on GitHub (https://github.com/opensafely/covid19-vaccine-coverage-tpp-emis).

### Patient and Public Involvement

Patients were not formally involved in developing this specific study design that was developed rapidly in the context of the rapid vaccine rollout during a global health emergency. We have developed a publicly available website https://opensafely.org/ through which we invite any patient or member of the public to contact us regarding this study or the broader OpenSAFELY project.

## Results

### COVID-19 vaccine coverage in JCVI priority groups

20,852,692 (36.0%) of patients registered at an EMIS or TPP practice received their first COVID-19 vaccine up to March 17th 2021 (Table 1, Figure 1). Of these, 275,205 were identified as living in a care home (group 1, 91.2% coverage of all care home residents), 2,422,476 were aged 80 and over (group 2, 94.7% of all over 80s not identified as living in a care home); and 1,865,927 were aged 75 −79 (group 3, 94.5% coverage). In later priority groups, 4,113,627 were identified as aged 70 − 74 or clinically extremely vulnerable (group 4, 87.7% coverage); 2,210,110 aged 65 - 69 (group 5, 89.4% coverage); and all other priority groups had a least 32.5% vaccination coverage. Among the youngest of the priority groups (50-54 and 55-59) some of those vaccinated to date are likely health and care workers. 3,012,051 patients, most likely health and care workers, were identified as being vaccinated but not assigned to one of the priority groups. Of all those vaccinated 12,080,194 (57.9%) received the *AstraZeneca-Oxford* vaccine as their first dose, 8,691,536 (41.7%) the *Pfizer-BioNTech* vaccine, with the remainder receiving a vaccine where the brand was not specified (n=80,962). 1,257,914 received a second dose (6.0%).

**Table 1.** Vaccination coverage among 95% of the population of registered patients in England, according to COVID-19 vaccination priority groups, at March 17th 2021. All ages are included.

| Priority Group | Vaccinated at 17th Mar (%) | Vaccinated at 17th Mar (n) | Population | Vaccinated at 10th Mar (%) | Vaccinated between 10th Mar and 17th Mar (%) |
| --- | --- | --- | --- | --- | --- |
| **1 - In care home** | 91.2% | 275,205 | 301,889 | 90.4% | 0.7% |
| **2 - 80+** | 94.7% | 2,422,476 | 2,558,906 | 94.5% | 0.2% |
| **3 - 75-79** | 94.7% | 1,865,927 | 1,970,234 | 94.5% | 0.2% |
| **4 - CEV or 70-74** | 87.7% | 4,113,627 | 4,690,203 | 86.7% | 1.0% |
| **5 - 65-69** | 89.4% | 2,210,110 | 2,472,862 | 87.6% | 1.7% |
| **6 - At risk** | 68.7% | 2,861,474 | 4,165,987 | 60.1% | 8.6% |
| **7 - 60-64** | 76.9% | 1,644,636 | 2,139,200 | 63.7% | 13.2% |
| **8 - 55-59** | 51.6% | 1,438,913 | 2,790,228 | 30.2% | 21.4% |
| **9 - 50-54** | 32.5% | 1,008,273 | 3,097,668 | 22.1% | 10.5% |
| **Other** | 8.9% | 3,012,051 | 33,743,290 | 8.0% | 1.0% |
| **Population** | 36.0% | 20,852,692 | 57,930,467 | 32.6% | 3.4% |

**Figure 1a.**
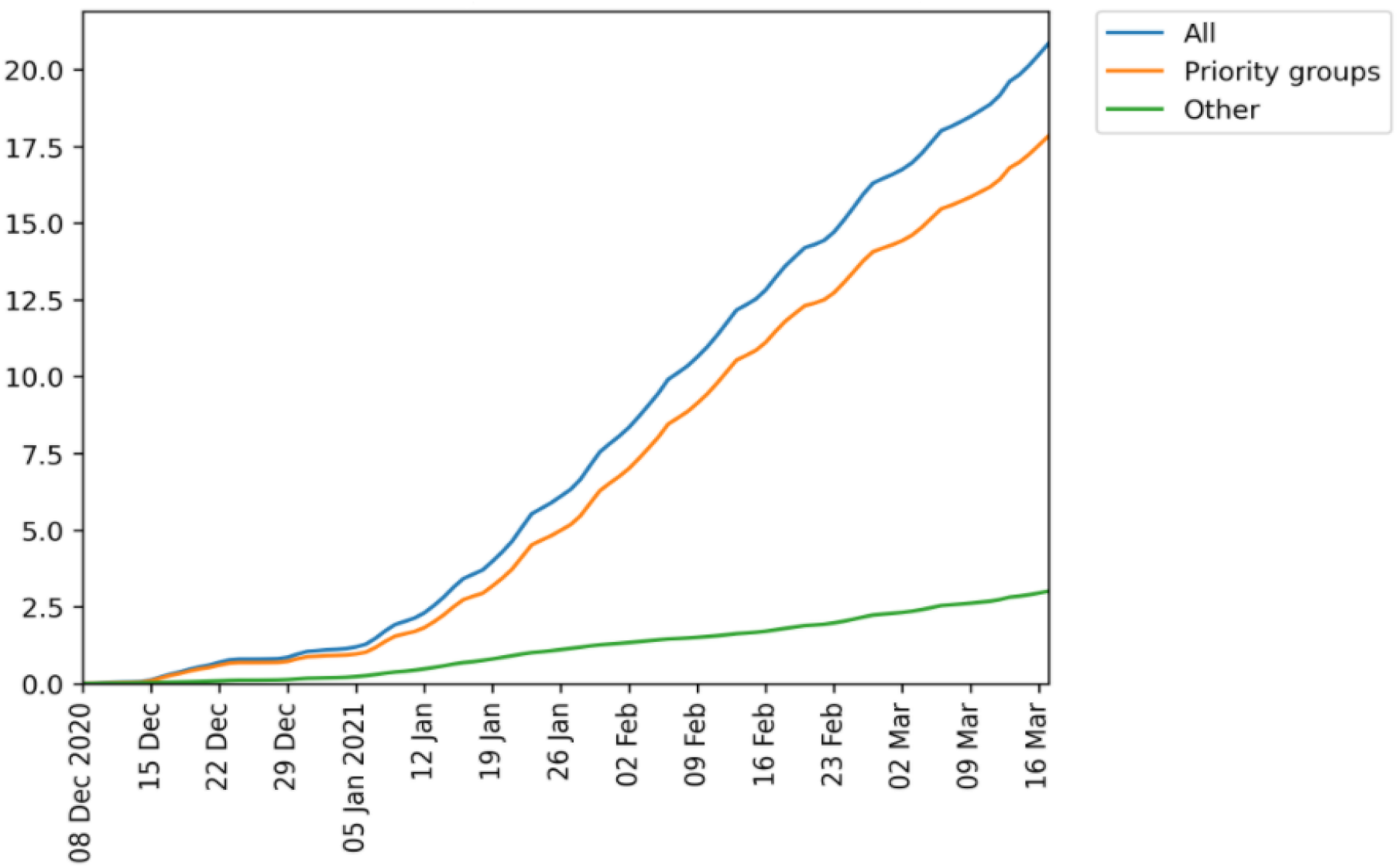
Cumulative daily total number of patients (millions) in England receiving COVID-19 at least one vaccination recorded in OpenSAFELY on March 17th 2021; across the whole population (All) and split according to whether patients were known to be part of a Priority Group or not (excluding occupational eligibility).

**Figure 1b.**
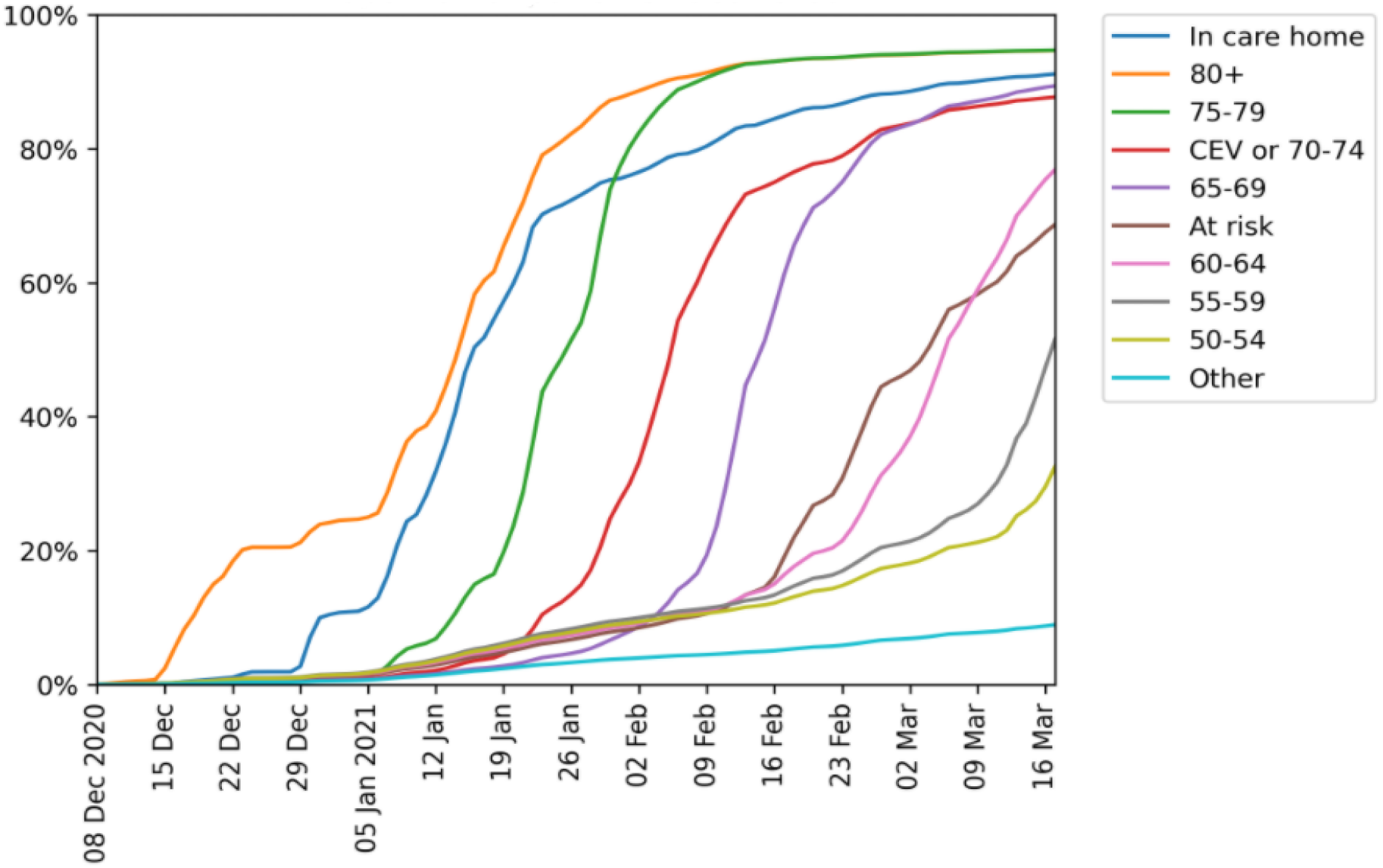
Cumulative daily proportion (%) of patients receiving at least one COVID vaccine dose by JCVI priority groups (excluding occupational eligibility) in England recorded in OpenSAFELY on March 17th 2021. CEV = Clinically Extremely Vulnerable.

### Key demographic and clinical characteristics of priority group 2

The substantially larger priority group 2 was offered vaccination alongside group 1. Priority group 2 included those aged 80 and over and not known to be living in a care home, and rapidly reached 90% coverage over the first eight weeks of the vaccination campaign, with a further increase of 4.7% over the remaining weeks of our study period. We show a breakdown of the proportion of patients vaccinated by March 17th in this group by various demographic and clinical categories in table 2 and figures 2 and 3. Vaccination was less common among those living in the most deprived postcode areas (90.7% in the most deprived quintile compared to 96.6% in the least deprived figure 2a). Across broad ethnic groups (figure 2b), the proportion vaccinated to date was highest among white ethnicity (96.2%); lowest among among black ethnicity (68.3%); 77.7%-84.6% among patients of mixed, other and South Asian ethnicities; and 94.1% among those with unknown ethnicity; ethnicity coding was 63.9% complete in this age group. Vaccination coverage in detailed ethnic vaccinations ranges from 96.7% in the “*White-British*” group to 59.8% in the A*frican: Back or Black British* group (table 2).

**Table 2.** COVID vaccinations among Priority group 2 (80+ population not resident in care homes) as at 17th Mar 2021, according to demographic and clinical features. Values <7 suppressed. Patient counts are rounded to the nearest 7.

|  |  | Vacc (%) at 17th Mar | Vacc (n) at 17th Mar | Population | Vacc (%) 10th Mar | Vacc between 10th Mar and 17th Mar (%) |
| --- | --- | --- | --- | --- | --- | --- |
| **Overall** | - | 94.7% | 2,422,476 | 2,558,906 | 94.5% | 0.2% |
| **Sex** | **Female** | 94.5% | 1,400,532 | 1,481,970 | 94.3% | 0.2% |
|  | **Male** | 94.9% | 1,021,944 | 1,076,936 | 94.8% | 0.1% |
| **Ethnicity** | **White - British** | 96.7% | 1,391,404 | 1,439,613 | 96.5% | 0.1% |
|  | **White - Irish** | 93.6% | 19,579 | 20,923 | 93.4% | 0.2% |
|  | **White - Any other White background** | 86.5% | 47,551 | 54,992 | 86.3% | 0.2% |
|  | **Mixed - White and Black Caribbean** | 75.0% | 2,779 | 3,703 | 74.5% | 0.6% |
|  | **Mixed - White and Black African** | 68.3% | 798 | 1,169 | 67.1% | 1.2% |
|  | **Mixed - White and Asian** | 86.2% | 1,134 | 1,316 | 85.6% | 0.5% |
|  | **Mixed - Any other mixed background** | 81.5% | 1,918 | 2,352 | 81.2% | 0.3% |
|  | **Asian or Asian British - Indian** | 89.9% | 29,456 | 32,753 | 89.7% | 0.2% |
|  | **Asian or Asian British - Pakistani** | 76.9% | 12,859 | 16,723 | 76.0% | 0.9% |
|  | **Asian or Asian British - Bangladeshi** | 81.3% | 3,969 | 4,879 | 80.5% | 0.9% |
|  | **Asian or Asian British - Any other Asian background** | 82.3% | 9,548 | 11,599 | 82.0% | 0.4% |
|  | **Black or Black British - Caribbean** | 71.3% | 16,681 | 23,387 | 70.6% | 0.7% |
|  | **Black or Black British - African** | 59.8% | 5,152 | 8,617 | 59.0% | 0.8% |
|  | **Black or Black British - Any other Black background** | 69.8% | 1,750 | 2,506 | 69.0% | 0.8% |
|  | **Other ethnic groups - Chinese** | 78.1% | 3,199 | 4,095 | 77.9% | 0.2% |
|  | **Other ethnic groups - Any other ethnic group** | 78.3% | 5,348 | 6,832 | 77.8% | 0.5% |
|  | **Patients with any other ethnicity code** | 95.4% | 377,566 | 395,654 | 95.3% | 0.2% |
|  | **Ethnicity not stated** | 92.9% | 15,344 | 16,513 | 92.7% | 0.2% |
|  | **Ethnicity not recorded** | 93.2% | 476,336 | 511,182 | 93.0% | 0.2% |
| **Ethnicity (broad categories)** | **White** | 96.2% | 1,458,548 | 1,515,535 | 96.1% | 0.1% |
|  | **Mixed** | 77.7% | 6,650 | 8,554 | 77.3% | 0.5% |
|  | **South Asian** | 84.6% | 55,846 | 65,975 | 84.2% | 0.5% |
|  | **Black** | 68.3% | 23,590 | 34,517 | 67.6% | 0.8% |
|  | **Other** | 78.3% | 8,561 | 10,934 | 77.9% | 0.4% |
|  | **Unknown** | 94.1% | 869,260 | 923,363 | 94.0% | 0.2% |
| **Index of Multiple Deprivation** | **1 (most deprived)** | 90.7% | 309,190 | 340,928 | 90.4% | 0.3% |
|  | **2** | 92.9% | 402,836 | 433,622 | 92.7% | 0.2% |
|  | **3** | 95.0% | 514,682 | 541,737 | 94.8% | 0.2% |
|  | **4** | 95.9% | 564,410 | 588,546 | 95.8% | 0.1% |
|  | **5 (least deprived)** | 96.6% | 612,731 | 634,340 | 96.5% | 0.1% |
|  | **Unknown** | 94.5% | 18,613 | 19,691 | 94.3% | 0.2% |
| **Clinical Risk Groups** | **Patients in Any Clinical Risk Group** | 95.7% | 1,766,590 | 1,846,677 | 95.5% | 0.2% |
|  | **Patients with Immunosuppression** | 96.6% | 107,184 | 110,999 | 96.4% | 0.2% |
|  | **Patients with CKD** | 95.9% | 750,820 | 782,705 | 95.8% | 0.2% |
|  | **Patients who have Chronic Respiratory Disease** | 96.0% | 294,413 | 306,810 | 95.8% | 0.2% |
|  | **Patients with Diabetes** | 94.8% | 492,982 | 519,862 | 94.6% | 0.2% |
|  | **Patients with Chronic Liver disease** | 95.4% | 44,618 | 46,746 | 95.2% | 0.2% |
|  | **Patients with CNS Disease (including Stroke/TIA)** | 95.4% | 408,730 | 428,400 | 95.2% | 0.2% |
|  | **Patients with Chronic heart disease** | 96.1% | 1,042,034 | 1,084,643 | 95.9% | 0.2% |
|  | **Patients with Asplenia or Dysfunction of the Spleen** | 96.8% | 19,950 | 20,615 | 96.7% | 0.1% |
|  | **Patients with Learning Disability** | 91.4% | 1,190 | 1,302 | 91.4% | 0.0% |
|  | **Patients with Severe Mental Health** | 89.5% | 15,582 | 17,409 | 89.1% | 0.4% |
| **Other Groups** | **Patients who are shielding (High Risk from COVID-19)** | 93.6% | 706,629 | 754,789 | 93.4% | 0.2% |
|  | **Patients with Morbid Obesity** | 93.8% | 32,837 | 35,014 | 93.5% | 0.3% |

**Figure 2.**
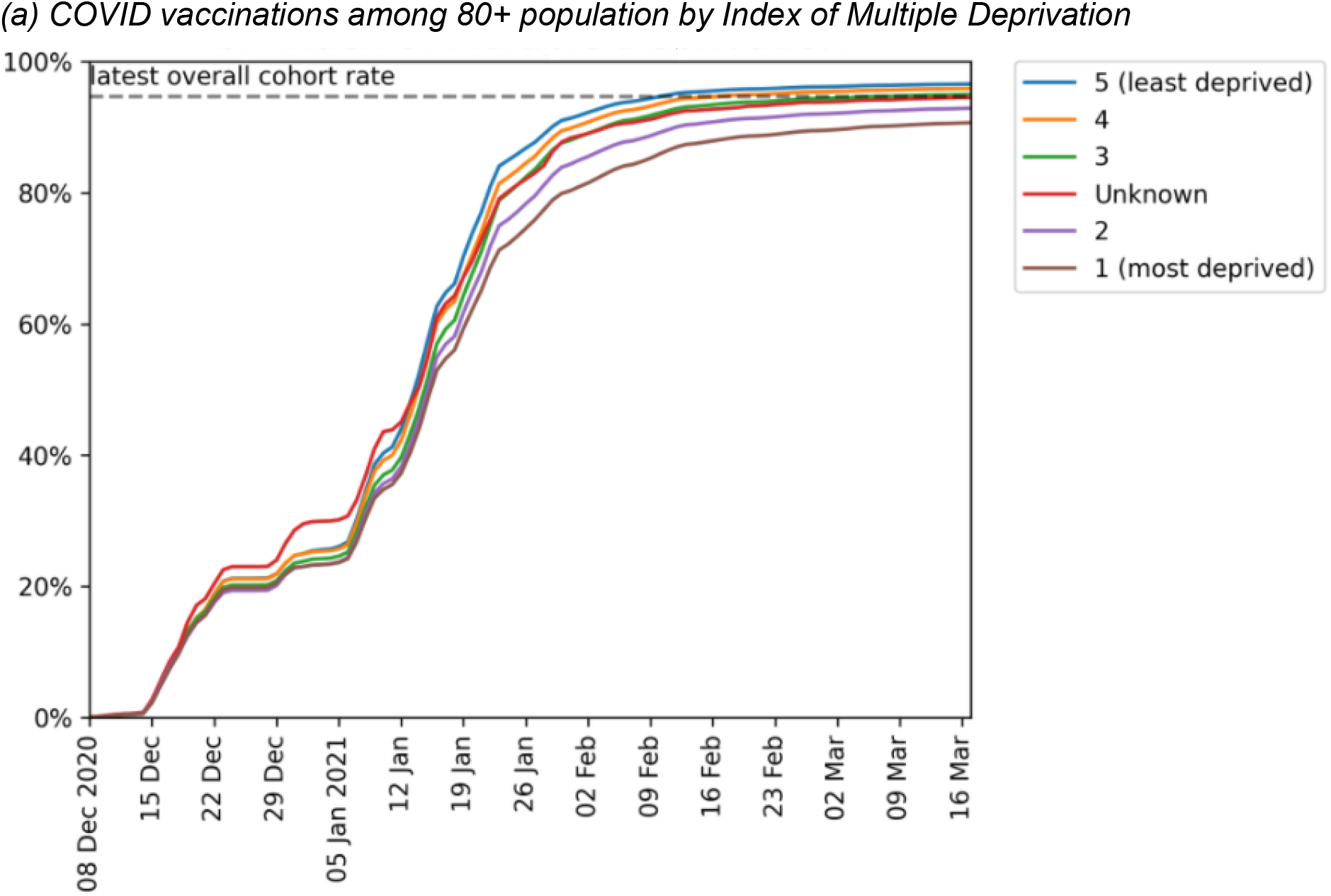
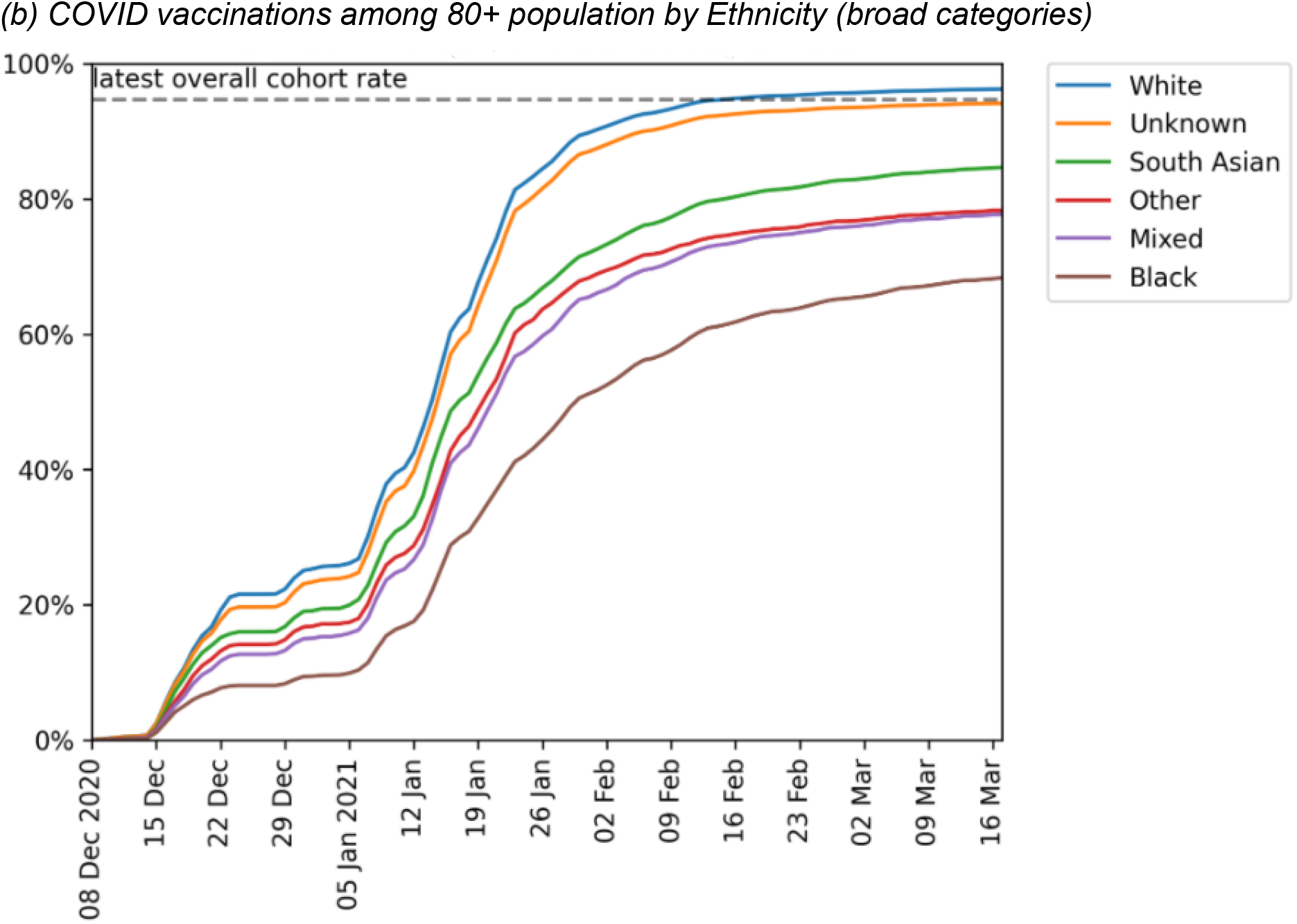
Cumulative trends in demographic features of the 80+ population receiving their first COVID vaccination.

**Figure 3.**
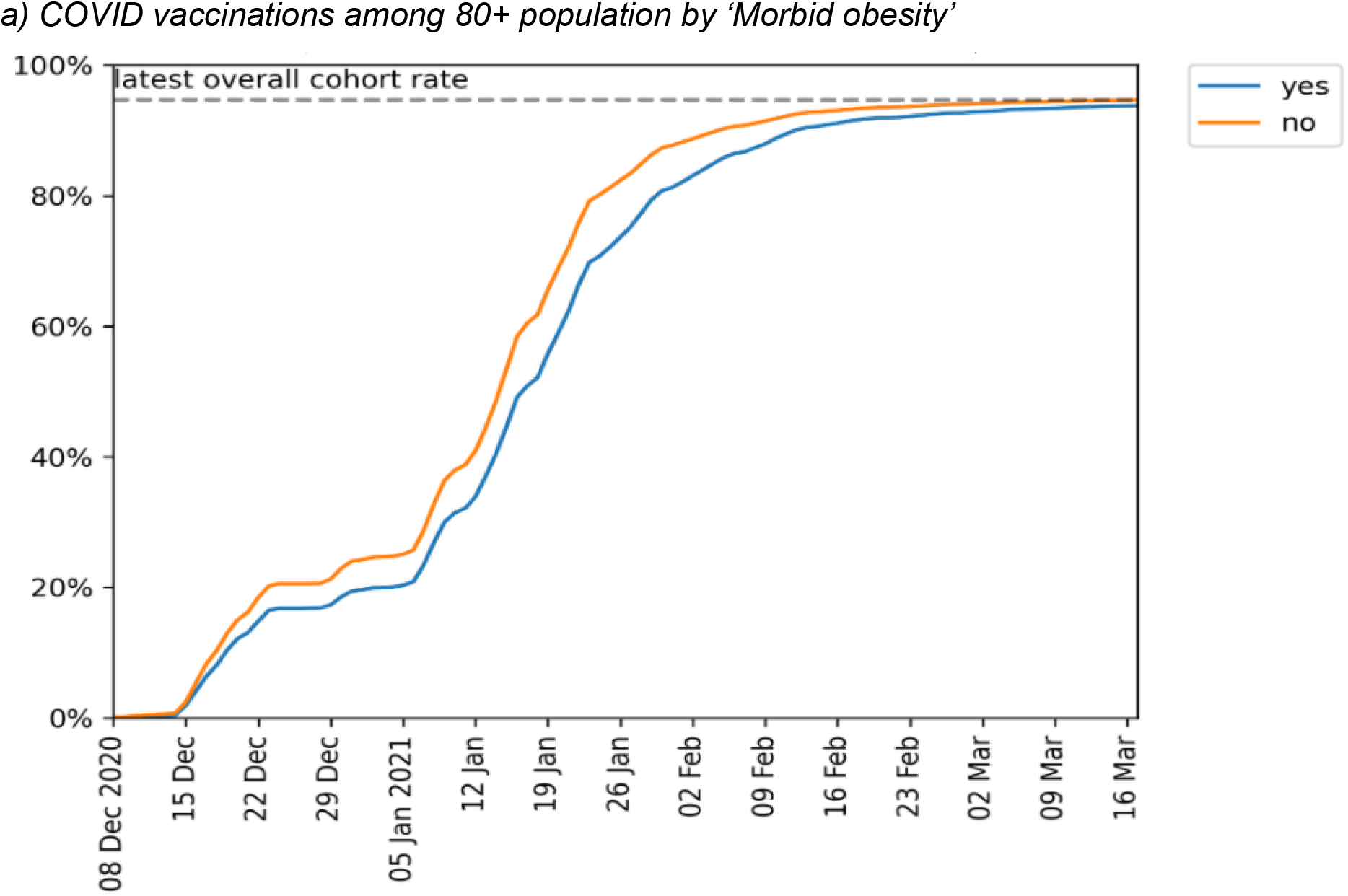
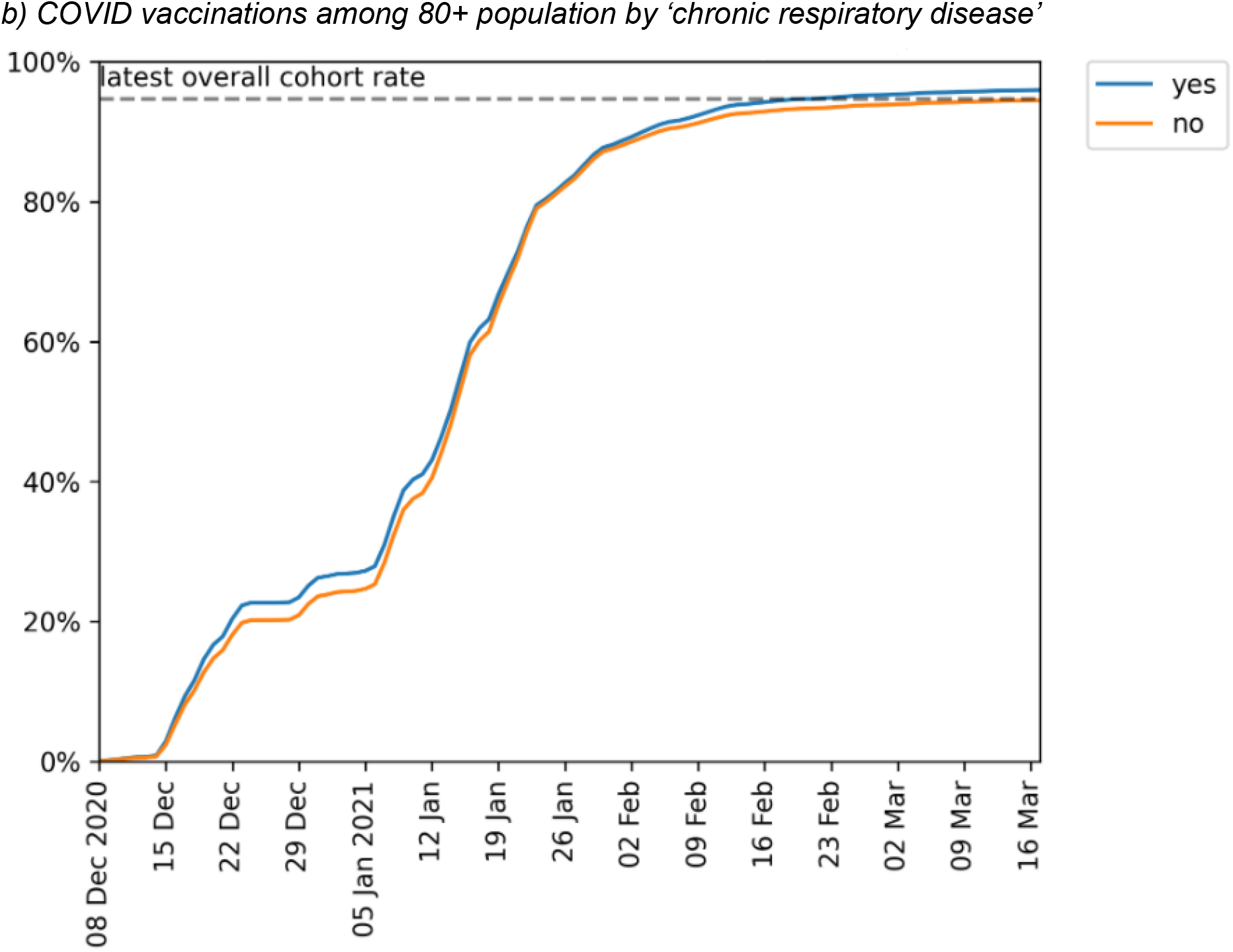
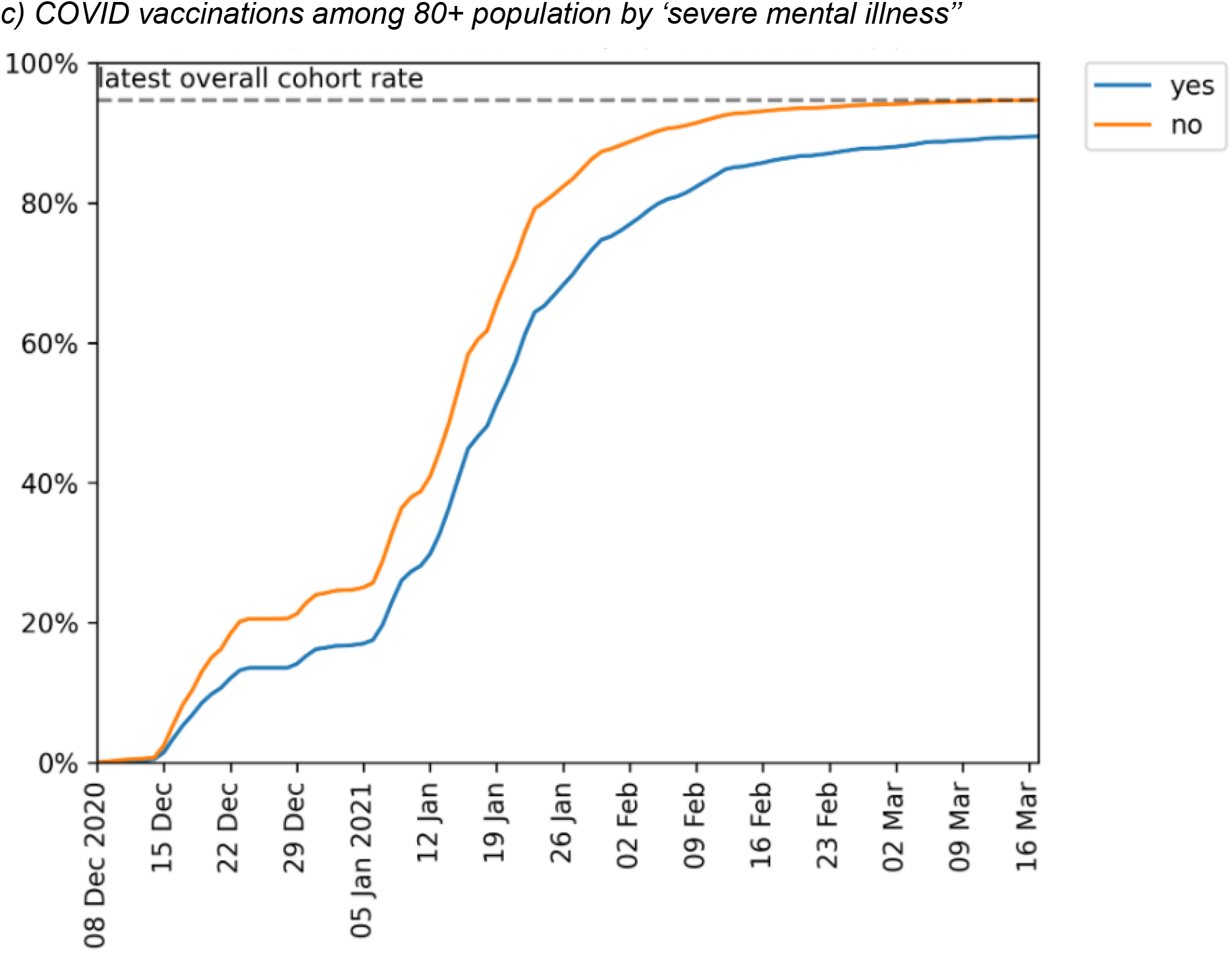
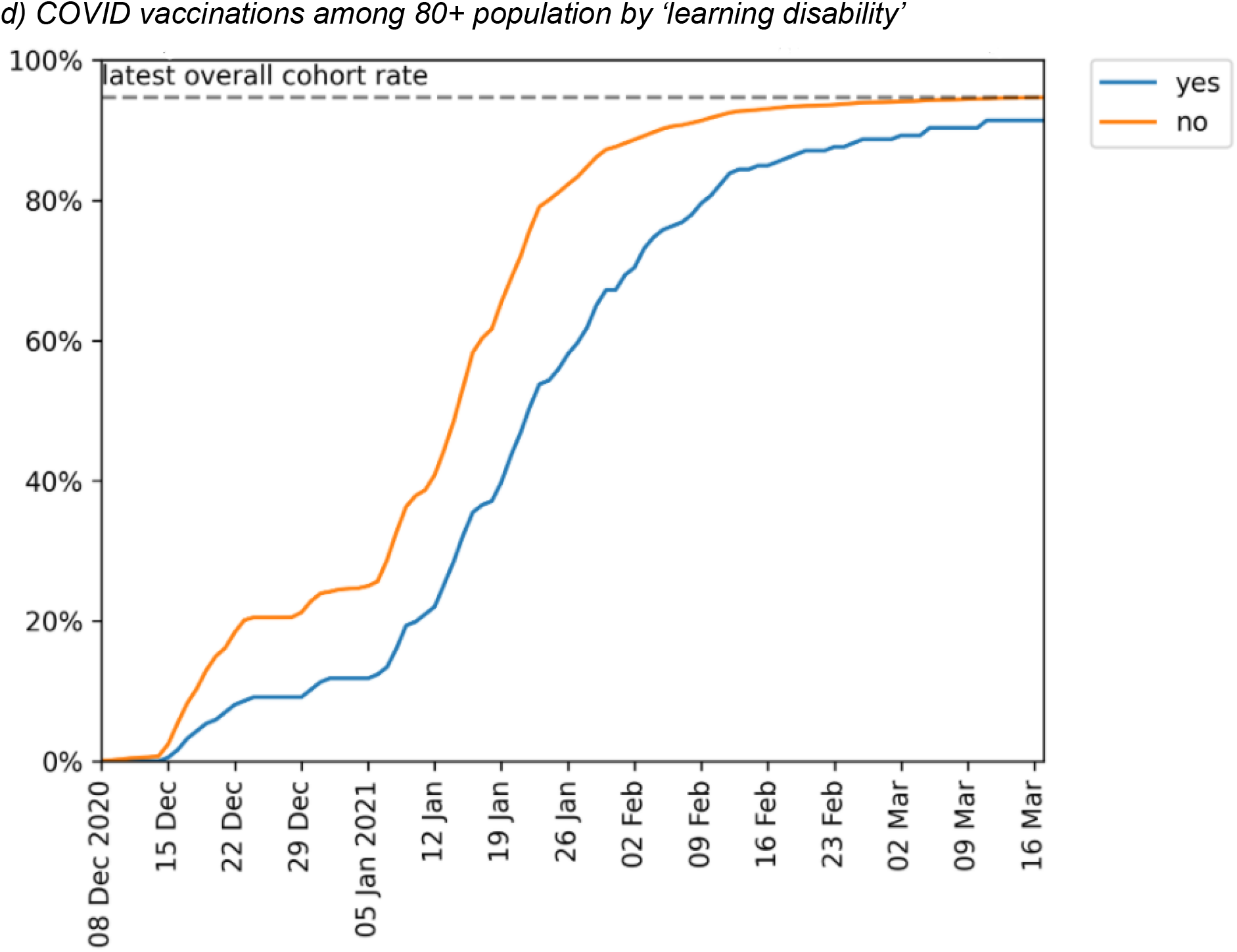
Cumulative trends in clinical characteristics of priority group 2 (aged 80+ and not resident in a care home) receiving their first COVID vaccination.

Vaccination coverage was initially lower among those with obesity (figure 3a); however this gap had largely resolved by mid-March 2021. Vaccination coverage was slightly higher among patients with other physical comorbidities such as chronic respiratory disease (figure 3b), cardiac disease, or chronic kidney disease. Vaccination coverage was substantially lower among those living with severe mental illness (89.5%, figure 3c) and learning disabilities (91.4%, figure 3d), with some improvements over time.

### Key demographic and clinical characteristics of other Priority groups

Detailed tables and charts of breakdowns for all other priority groups are available on our GitHub repository [14]. The differences by demographic and clinical features observed in priority group 2 were broadly reflected in other priority groups. Key exceptions were that the lower priority groups, not yet widely vaccinated, had a different pattern of discrepancy by ethnicity, with the South Asian population most vaccinated. This pattern was also seen in several other groups during early stages of the campaign, prior to widespread targeting of individual groups, such as the 70-74 and 75-79 groups. We also note recent improvements in the vaccination coverage of the Bangledeshi community, where an acceleration with respect to other ethnic groups was seen from around the 16th February, particularly noticeable in priority groups 4, 6, 7, 8 and 9.

## Discussion

### Summary

The NHS in England has rapidly responded to the availability of COVID-19 vaccines and administered a substantial number of doses in the first 100 days of the vaccination campaign. In our study 20,852,692 patients (36.0% of patients registered in 97% of GP practices in England) received at least one vaccine dose by March 17th, including 94.7% of eligible patients aged 80 and over (priority group 2). However, ethnic minorities in priority group 2 were substantially less likely to be vaccinated, and those living in more socioeconomically deprived areas generally had lower vaccine coverage. Similarly these patterns were broadly observed across the majority of priority groups. Furthermore, in priority group 2 in those aged 80 or over, patients with pre-existing medical conditions were equally likely, or more likely, to have received a vaccine, across all groups of pre-existing medical problems, with two exceptions: vaccination was lower among patients living with severe mental illnesses and learning disabilities. 3,012,051 people who received a vaccination, most likely health and care workers, were not identified as part of any priority group. Usage was split between two vaccine brands, *Pfizer-BioNtech* (41.7% n=8,691,536) and *AstraZeneca-Oxford* (57.9%, n=12,080,194). 6.0% (n=1,257,914) received a second dose of the vaccine.

### Strengths and weaknesses

The key strengths of this study are the scale, detail, completeness and timeliness of the underlying raw EHR data. Our analysis was executed across the full dataset of all raw, pseudonymised, single-event-level clinical events for 57.9 million patients registered at all NHS GP practices in England using EMIS and TPP software; this includes data on all tests, treatments, diagnostic events, and other salient clinical and demographic information. This was achieved by developing and deploying data management and data analysis software inside the EHR vendors’ infrastructure, where the patient data already resides. As a consequence of this, OpenSAFELY can deliver insights into health service activity and clinical outcomes in near-real time: raw data can be processed into a completed analysis within days of a clinical event being entered into the patient’s record. Another key strength is that we identified all eligible patients in each JCVI priority group by directly implementing the full official SNOMED-CT codelists and logic for the national PRIMIS *COVID-19 Vaccination Uptake Reporting Specification*, thus ensuring that our cohorts are perfectly in line with national procedures and GP expectations.

We recognise some limitations to our analysis. Our population, though extremely large, may not be fully representative of the full eligible population: it does not include individuals not registered with a general practice; or the 4% of patients registered at practices not using TPP and EMIS. Primary care records, whilst detailed and longitudinal, can be incomplete on certain patient characteristics. Occupation is generally not available in the health record so we were unable to assess the eligibility in priority groups 1 or 2 where this is based on occupation. This means that we could not determine the appropriateness of vaccination for those in the “other” group. For patients aged 80 and over, 36.2% and 0.8% had missing ethnicity or IMD information respectively. In our weekly COVID vaccination coverage report we use a different ethnicity codelist and additional sources of data within OpenSAFELY-TPP to reduce missing data to less than 10% for ethnicity [11]. We will implement this method within OpenSAFELY-EMIS shortly. As we have carried out a federated analysis across two EHR vendors systems, it is possible that a very small number of patient records are duplicated; we are currently exchanging a pseudonymised list of NHS numbers between the two settings to eradicate this issue. Our ascertainment of vaccination status relies on the vaccination administration electronic message being successfully received into the primary care record; while our numbers are consistent with national figures, we are exploring methods to also cross-validate this against other sources of person-level vaccination data, broken down by vaccination site type. Lastly, there is currently no well-validated person-level data to identify individuals resident in a care home: this is a limitation for all UK healthcare database studies: the method used for identifying care home residents in this analysis - a clinical code as detailed by the national reporting specification - will lead to under-ascertainment [15]. We are launching a programme of work, in collaboration with the UK health data science community, to describe and validate the best methods for identifying current care home residents, in order to produce a better understanding of their health outcomes.

### Findings in Context

The UK has already administered 41.65 vaccines per 100 people, one of the fastest vaccination programmes in the world [16]. NHS England publish reports counts of vaccines delivered using vaccination data extracted from the NIMS and report that 21,886,125 first dose vaccinations were administered at March 17th [17]: this is in line with our finding of 20,852,692 patients vaccinated in OpenSAFELY data. It is reasonable to expect marginal differences in the count of patients vaccinated, and the proportions vaccinated, between different summary reports from different analytic teams, due to minor differences in the speeds of data flow, and in the ascertainment of denominators. Our total OpenSAFELY population figure is 57.9 million, which is approximately 95% of the NHS Digital estimate for the number of people registered with an NHS GP (60.65million) [18]. We note that this figure is higher than the latest ONS estimated population of England (56.2million)[19]: the difference between ONS population estimates and NHS registered populations is a well recognised issue and may be caused by over-counting at GP practices, differences in definition and under-counting by the ONS, or a combination of all three [20,21].

To our knowledge this manuscript, an update of our January 27th preprint covering only OpenSAFELY-TPP patients (40% of the population), is the first study to describe in detail the demographic and clinical features of those who have been vaccinated by the NHS England COVID-19 vaccination campaign; and the only study to report on variation in vaccination by fine-grained clinical characteristics, because OpenSAFELY can provide detailed information about the demographics and clinical conditions of those vaccinated from each patients’ full pseudonymised EHR, which is not available within NIMS. Our finding of lower vaccination coverage in black and asian groups is concerning: it is also consistent with previous research on variation in vaccine coverage between ethnic groups during other vaccination campaigns historically [22–24], and with survey data on intention to accept the COVID-19 vaccine. [25–28].

### Policy Implications and Interpretation

The reasons underpinning variation in COVID-19 vaccination coverage are not yet understood, and information presented here should not be misinterpreted as a criticism of the rapidly established NHS vaccination campaign. Further research is needed to understand and address the observed lower vaccination coverage among patients from more deprived areas, and the striking disparity between ethnic groups. Our initial preprint on January 27th and our regular updates [11,29] have received substantial media coverage particularly with regards to the differences in vaccinations between different ethnic communities [30–33]. The NHS, government and communities themselves have introduced targeted activities to address the gap including vaccination at places of worship [34], webinars led by community leaders to tackle misinformation [35], and targeted funding for groups with remit for tackling any health inequalities [36,37]. We note the accelerated increase in the Bangladeshi community from mid-February, which may represent targeted action by a community group and/or a local NHS organisation. The regular OpenSAFELY vaccination coverage reports can support assessment of the success of these activities in increasing vaccination coverage.

It is reassuring to see that those with a previous history of various medical problems are being vaccinated at the same rate as other patients: in particular it is reassuring to see no evidence that the vaccine programme is currently missing those with serious physical health problems who are at highest risk of death from COVID-19. The lower vaccination coverage among patients living with severe mental illness and learning disabilities is concerning: this may reflect challenges around access, including for those currently living in institutional settings. However in recent weeks there is evidence that the vaccination gap is narrowing in these groups (figure 3). In late February the JCVI recommended expanded vaccine access for all people on the GP learning disability register as well as adults with other related conditions, including cerebral palsy. This update of the previous advice was based on OpenSAFELY analysis showing a higher risk of mortality in those learning disabilities [38] and the JCVI anticipated that an additional 150,000 people would receive the vaccine sooner as a result of this advice [39].

We note that all findings in this paper are from the first few months of a major national vaccination programme. Very substantial changes in coverage among different groups are to be expected over the coming months. We are openly sharing weekly vaccine coverage reports to assist in monitoring and targeting vaccine initiatives along with machine readable outputs for re-use in different formats. These updates are available at www.opensafely.org/covid-vaccine-coverage. More broadly, the UK has an unusually large volume of very detailed longitudinal patient data, especially through primary care. We believe the UK has a responsibility to the global community to ensure that this data is used to inform response to COVID-19 pandemic, in a timely manner, whilst maintaining the security of individual health records and ensuring the full transparency of all actions on the data to build public trust. To this end, codelists and code for data management and data analysis can only be executed on OpenSAFELY after first being made available at GitHub.com/openSAFELY prior to execution; this is then shared publicly under open licenses for review and re-use either at, or before, the time when results are reported.

### Summary

The NHS in England has rapidly deployed a mass vaccination campaign. Targeted activity may be needed to address lower vaccination coverage observed among certain key groups: ethnic minorities, those living in areas of higher deprivation, and individuals living with severe mental illness or learning disabilities. Live data monitoring is likely to help support those on the front line making complex operational decisions around vaccine roll-out.

## Data Availability

Data management and analysis was performed using the OpenSAFELY software libraries and Jupyter notebooks, both implemented using Python 3. More details are available in the Supplementary Materials. This is the first analysis delivered using federated analysis through the OpenSAFELY platform: codelists and code for data management and data analysis were specified once using the OpenSAFELY tools; then transmitted securely from the OpenSAFELY jobs server to the OpenSAFELY-TPP platform within TPPs secure environment, and separately to the OpenSAFELY-EMIS platform within EMISs secure environment, where they were each executed separately against local patient data; summary results were then reviewed for disclosiveness, released, and combined for the final outputs. All code for the OpenSAFELY platform for data management, analysis and secure code execution is shared for review and re-use under open licenses at GitHub.com/OpenSAFELY. All code for data management and analysis for this paper is shared for scientific review and re-use under open licenses on GitHub (https://github.com/opensafely/covid19-vaccine-coverage-tpp-emis). 

https://github.com/opensafely/covid19-vaccine-coverage-tpp-emis

## Administrative

### Funding

This work was supported by the Medical Research Council MR/V015737/1 and the Longitudinal Health and Wellbeing strand of the National Core Studies programme. EMIS and TPP provided technical expertise and infrastructure within their data environments *pro bono* in the context of a national emergency. The OpenSAFELY software platform is supported by a Wellcome Discretionary Award. BG’s work on clinical informatics is supported by the NIHR Oxford Biomedical Research Centre and the NIHR Applied Research Collaboration Oxford and Thames Valley. Funders had no role in the study design, collection, analysis, and interpretation of data; in the writing of the report; and in the decision to submit the article for publication. The views expressed are those of the authors and not necessarily those of the NIHR, NHS England, Public Health England or the Department of Health and Social Care.

### Information governance and ethical approval

NHS England is the data controller; EMIS and TPP are the data processors; and the key researchers on OpenSAFELY are acting on behalf of NHS England. This implementation of OpenSAFELY is hosted within the EMIS and TPP environments which are accredited to the ISO 27001 information security standard and are NHS IG Toolkit compliant;[40,41] patient data has been pseudonymised for analysis and linkage using industry standard cryptographic hashing techniques; all pseudonymised datasets transmitted for linkage onto OpenSAFELY are encrypted; access to the platform is via a virtual private network (VPN) connection, restricted to a small group of researchers; the researchers hold contracts with NHS England and only access the platform to initiate database queries and statistical models; all database activity is logged; only aggregate statistical outputs leave the platform environment following best practice for anonymisation of results such as statistical disclosure control for low cell counts.[42] The OpenSAFELY research platform adheres to the obligations of the UK General Data Protection Regulation (GDPR) and the Data Protection Act 2018. In March 2020, the Secretary of State for Health and Social Care used powers under the UK Health Service (Control of Patient Information) Regulations 2002 (COPI) to require organisations to process confidential patient information for the purposes of protecting public health, providing healthcare services to the public and monitoring and managing the COVID-19 outbreak and incidents of exposure; this sets aside the requirement for patient consent.[43] Taken together, these provide the legal bases to link patient datasets on the OpenSAFELY platform. GP practices, from which the primary care data are obtained, are required to share relevant health information to support the public health response to the pandemic, and have been informed of the OpenSAFELY analytics platform. This study was approved by the Health Research Authority (REC reference 20/LO/0651) and by the LSHTM Ethics Board (reference 21863).

### Guarantor

BG/LS are guarantors of the OpenSAFELY project.

## Acknowledgements

We are very grateful for all the support received from the EMIS and TPP Technical Operations team throughout this work, and for generous assistance from the information governance and database teams at NHS England / NHSX.

